# What is the relation between major depressive disorder and amygdala reactivity?

**DOI:** 10.64898/2025.12.24.25342967

**Authors:** Yu Hao, Colin Xu, Hyeokmoon Kweon, Martha J. Farah

## Abstract

The present study focuses on the role of amygdala reactivity to negative facial expressions in major depressive disorder (MDD). A number of studies have found amygdala hyperreactivity in depressed patients compared with control subjects. This has been interpreted in terms of a negative depressive bias in attention and memory, given the amygdala’s role in attending to and remembering negatively valenced stimuli. However, failure to find amygdala hyperreactivity in depression is not uncommon, and a recent failure to replicate the effect with an extremely well-powered analysis of UK Biobank data led many to conclude that amygdala reactivity plays no role in depression. In the present study, the same UK Biobank sample is used to evaluate an alternative hypothesis about the relation between MDD and amygdala reactivity, namely that people who are *vulnerable* to MDD, whether or not they are currently depressed, will show elevated amygdala activity. Depression history was used as a proxy for vulnerability and regression analyses assessed the strength of the relation between three MDD history measures and amygdala reactivity. Two of three assessments of depression history showed highly significant relations with amygdala reactivity, the third showing a combination of significant and nonsignificant trends depending on the analysis.

## Introduction

In a classic fMRI study by Hariri and colleagues (2002), participants performed a simple task in which they matched faces expressing negative emotions or matched geometric shapes. Consistent with the role of the amygdala in processing emotion, the emotional faces evoked more amygdala activity than the shapes. Subsequent research found that the “Hariri effect” is larger in depression (e.g., Fitzgerald et al., 2008). This elevated response is of interest because it suggests a neural bias in depression toward attending to or remembering negatively valenced stimuli (Disner et al., 2011).

However, as the field became more aware of the risk of false positives, particularly in the imaging literature with its flexibility of analytic approach and often small and medium samples (Botvinik-Nezer et al, 2020; Button et al., 2013), researchers have cast a more skeptical eye on reports of elevated amygdala activity in depression. A number of failures to replicate these studies have entered the literature, the best known of which comes from Tamm and colleagues (2022), who assessed amygdala reactivity in depression with data from approximately 30,000 middle-aged and older individuals in the UK Biobank. Despite the large sample, there was no relation between depression score and amygdala reactivity. This finding cast doubt on a role for amygdala reactivity in MDD and has been cited as decisive evidence on the matter (Fox & Shackman, 2024; Nour et al., 2022).

Using the same large and well-characterised sample as Tamm et al (2022), the analyses presented here were designed to assess the relation between amygdala reactivity and the trait of depression vulnerability, rather than the state of depression. Studies comparing state depression and euthymia, such as Tamm et al’s, may not show the effects of vulnerability because vulnerable individuals may not be depressed on the day of scanning. Given the presumed role of amygdala reactivity in attention to and memory for negatively valenced emotional stimuli (Phelps, 2006), one might expect heightened amygdala reactivity to indicate a negative bias, thereby predisposing to depression.

## Methods

### Participants

The sample was UK Biobank participants who attended the first imaging visit and who were free of the following neurological conditions: neurodegenerative disease, stroke, head injury, or epilepsy, resulting in n = 31,191. They were 63 years old (SD=7.62) on average at the time of scanning, and 47% female. Different numbers of these participants had different measures of depression history available (see Table 1 for resulting sample sizes).

**TABLE 1.** Caption: Different measures of lifetime depression history, described in the Methods, in relation to amygdala reactivity in the Hariri task. Significant contrasts bolded. *Key*: *\* p < 0*.*05, ** p < 0*.*01, *** p < 0*.*001*.

Part A
| **DEPRESSION MEASURE** | **NEVER DEPRESSED (reference group)** | **DEPRESSION - LIFETIME EVER** |
| --- | --- | --- |
| History of cardinal symptoms for at least 2 weeks reported to nurse at imaging visit<br>Total N = 26,582 | N = 14,095 (53.0%) | N = 12,487 (47.0%)<br>univariate:<br>**$\beta=0.0505$\*\*\***<br>**SE=0.0123**<br>**p<0.0001**<br>with covariates:<br>**$\beta= 0.0353$\*\*\***<br>**SE=0.0125**<br>**p=0.0047** |

Part B
| **DEPRESSION MEASURE** | **NEVER DEPRESSED (reference group)** | **DEPRESSION - LIFETIME EVER** |
| --- | --- | --- |
| Criteria described by Smith et al. (2013) reported to nurse at Imaging visit<br>Total N = 19,991 | N = 14,095 (70.5%) | N = 5,896 (29.5%)<br>univariate:<br>**$\beta= 0.0708$\*\*\***<br>**SE=0.0156**<br>**p<0.0001**<br>with covariates:<br>**$\beta= 0.0487$\*\***<br>**SE=0.0160**<br>**p=0.0023** |

Part C
| **DEPRESSION MEASURE** | **NEVER DEPRESSED (reference group)** | **DEPRESSION - LIFETIME EVER** |
| --- | --- | --- |
| CIDI-sf plus major role impairment as described by Cai et al. (2020); Self-administered online follow-up<br>Total N = 12,726 | N = 9,472 (74.4%)<br>Reference Group | N = 3,254 (25.6%)<br>univariate:<br>**$\beta= 0.0405$\***<br>**SE=0.0199**<br>**p=0.0421**<br>with covariates:<br>$\beta= 0.0238$<br>SE=0.0206<br>p=0.2470 |

### Measures of depression history

Burcusa and Iacono’s (2007) systematic review indicated that depression history is the best predictor of vulnerability to future depression, even after accounting for stress, neuroticism and family history, and therefore it was used as a proxy for depression vulnerability. We are agnostic as to whether this vulnerability is innate, or is acquired through the effects of previous episodes of depression (“scarring”). Vulnerable individuals will normally experience their first episode of depression by midlife, which makes depression history an especially useful proxy for vulnerability in this mid-to-late life sample.

Focusing on the depression history measures that have been reviewed and compared in recent UK Biobank studies (Cai et al., 2020; Harris et al., 2020), three were used for the present analysis. The measures of depression history used here differ from one another in several ways, including the quantity and nature of the measures (fewer or more symptoms, or additional indicators such as help-seeking), and the method by which answers were elicited (interview by research nurse on-site or online questionnaire that participants self-administered elsewhere. The sample sizes for which these measures were available differed, with the largest sample more than twice the smallest. Finally, the measures differed in time of measurement relative to scanning.

The first measure was the report, elicited by a project nurse, of past cardinal symptoms of MDD (depressed or anhedonic mood, Fields 4598, 4631) lasting at least two weeks (Fields 4609 and 5375). As shown in Table 1, this measure was available for over 26,000 participants from the imaging visit.

The second was a composite measure developed by Smith et al. (2013) based on reports, elicited by a nurse, of at least one cardinal symptom lasting at least two weeks (same fields as above), along with past consultation with a physician for this problem (Field 2090). Treatment by a psychiatrist (Field 2100) rather than a general practitioner was used as a measure of severity, on the assumption that GPs would generally refer only severely depressed patients to a psychiatrist. Number of past episodes was also recorded. These data were collected at the imaging visit. As shown in Table 1, just under 20,000 participants had these data available.

The third measure was adapted from the genetics study of Cai et al. (2020). It is based on the Composite International Diagnostic Interview – Short Form (Kessler et al., 1998), which probes the two cardinal symptoms of depression (sadness and anhedonia) and, contingent on a cardinal symptom being reported, five additional DSM Major Depressive Disorder symptoms. Additionally participants were required to report “a lot of” role impairment. The number of previous episodes was also queried and recorded. In contrast to the other two measures, participants self-reported this information online, outside of a visit to one of the testing centers. The date of data collection differed from the imaging visit by as much as 3 yr in either direction. As shown in Table 1, these data were available for almost 13,000 participants.

### Imaging task and statistical analyses

Amygdala reactivity was assessed using the UK Biobank’s implementation of the Hariri task at the first imaging visit (Alfaro-Almagro et al., 2018). We used the median BOLD effect for the faces-shapes contrast (Field 25052) as the dependent variable. Analyses consisted of linear regression comparing amygdala reactivity in participants with a history of depression and never-depressed participants. Additional analyses were undertaken to compare more specific types of depression history to no history, and are reported in the Supplementary Materials. Results are reported with and without the covariates used by Tamm et al (2022): sex, age, BMI, education and neighborhood deprivation.

## Results

Table 1 displays a summary of the results of the analyses just described. As shown in Part A, the report of at least one cardinal symptom of depression for at least two weeks was related to amygdala reactivity. This was true in both the univariate analysis and the multivariate analysis using covariates. Part B displays the results based on the criteria of Smith et al (2013). Those with depression histories showed greater amygdala reactivity compared to the never-depressed, with and without covariates. Finally, in Part C depression history by the criteria of Cai et al (2020) shows an association with amygdala reactivity by univariate analysis but not in the presence of covariates.

Analyses including recurrence and severity were also carried out, with results as shown in Supplementary Table S1. Statistical significance persisted after disaggregation into smaller groups using the Smith et al. (2013) classifications, but not using Cai et al. (2020)’s classifications.

## Discussion

What do these findings tell us about the role of amygdala reactivity in depression? We tested the hypothesis that a heightened Hariri effect exists and reflects depression vulnerability, using three measures of depression history available in the UK Biobank as a proxy for vulnerability. Two of three depression history measures provided clear support for this hypothesis, while the third (Cai’s adaptation of the CIDI-sf) yielded weaker support, showing significance in a univariate analysis and a nonsignificant trend when the five covariates were included. There is increasing awareness of the imperfect overlap in classifications of MDD using different criteria due to different weighting of characteristics across measures of depression (Fried, 2017; Huang et al., 2024). The smaller sample and the at-home self-administration may also contribute to the inconsistency between the results. Although the Smith criteria feature fewer questions than Cai’s, it should be noted that the Smith criteria are different but perhaps no less informative than the CIDI-sf-based criteria, in that being referred to a psychiatrist packs a number of physician observations into a single item.

The role of amygdala reactivity in vulnerability to depression could be questioned on the grounds that the effect sizes found here are very small, with standardized βs between 0.05 and 0.07 (univariate) and between 0.04 and 0.05 (multivariate). We grant this important point, but we also note that a bias in perception, attention and memory toward negative emotions would not be a “one-shot” occurrence, but rather a constant and ongoing bias. A small effect size operating many times daily may, like a river gradually eroding rock, result in large effects over time.

## Conclusion

MDD is likely to have many contributing causes. We suggest that one such cause may be the predisposition to attend to and remember negatively valenced events, consistent with elevated amygdala reactivity to negative facial expressions. If true, an enhanced Hariri effect may provide clues to the nature of depression vulnerability and might possibly mark distinctions among different kinds of depression, emerging from different pathways and responding to different preventive measures or treatments (Brotman et al., 2024; Phillips et al., 2015).

## Data Availability

All data produced are available online at UK biobank.
https://www.ukbiobank.ac.uk

## Acknowledgements

This research was carried out under the auspices of the Brain Imaging and Genetics in Behavioral Research Consortium (https://big-bear-research.org/), using UK Biobank resources under application 11425. We thank the School of Arts and Sciences at the University of Pennsylvania for funding. We thank Rob DeRubeis and Lois Gelfand for their expert commentary and feedback, and Philipp Koellinger and Gideon Nave for their advice on UK Biobank navigation and this research. The results were presented at the *Society for Neuroscience* meeting in Washington DC in 2023.

---

**TABLE S1.**
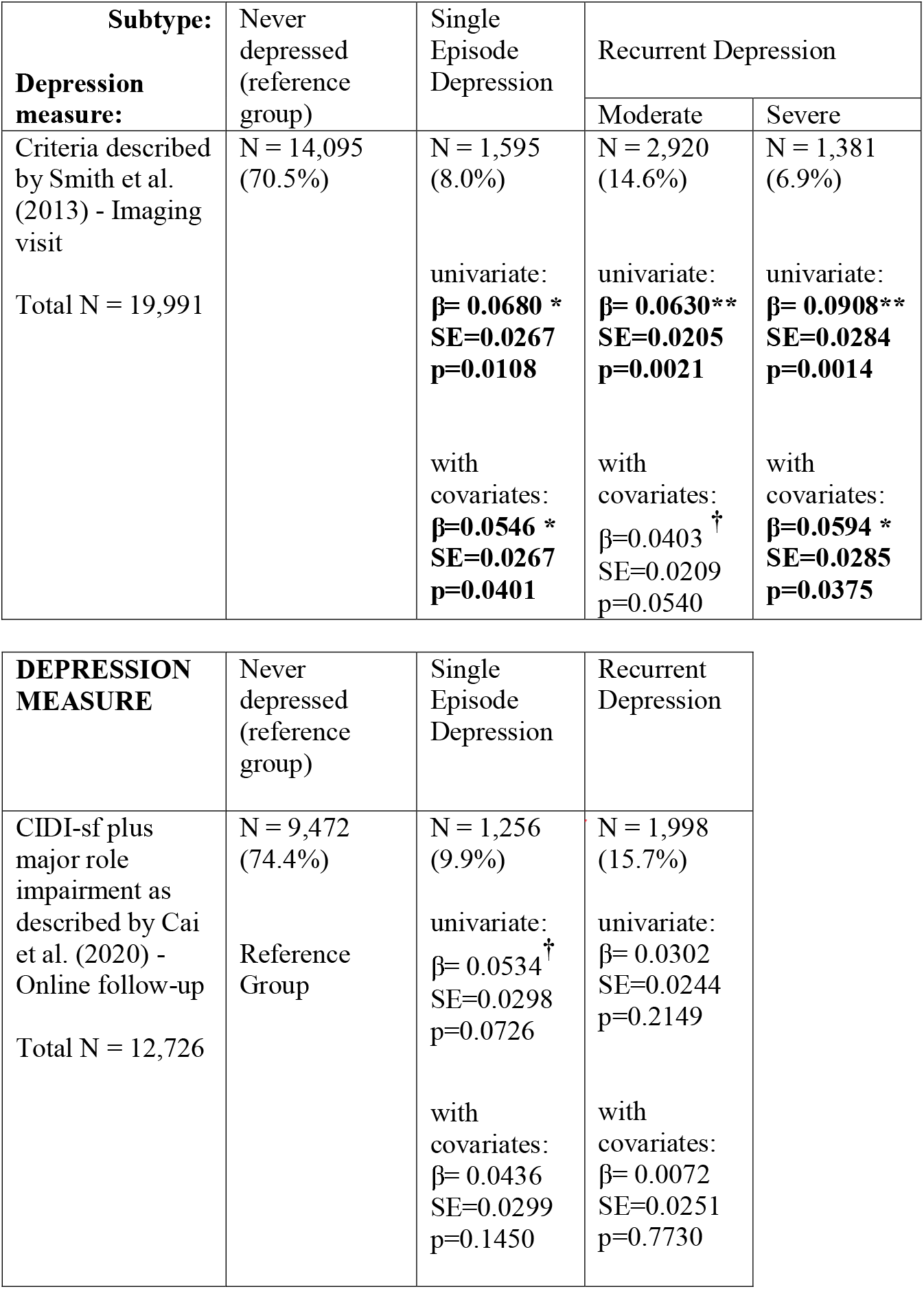
Caption: Disaggregation the depression history classifications of the different measures of lifetime depression history that include recurrence and severity, in relation to amygdala reactivity in the Hariri task. Each column represents one regression equation contrasting the respective group against the Never-depressed group. **†** Significant relations bolded. *Key*: *0*.*1 < p > 0*.*05, * p < 0*.*05, ** p < 0*.*01, *** p < 0*.*001*

| Subtype:<br>Depression<br>measure: | Never<br>depressed<br>(reference<br>group) | Single<br>Episode<br>Depression | Recurrent Depression |  |
| --- | --- | --- | --- | --- |
|  |  |  | Moderate | Severe |
| Criteria described<br>by Smith et al.<br>(2013) - Imaging<br>visit<br>Total N = 19,991 | N = 14,095<br>(70.5%) | N = 1,595<br>(8.0%)<br>univariate:<br>**$\beta = 0.0680$ \***<br>**SE=0.0267**<br>**p=0.0108**<br>with<br>covariates:<br>**$\beta = 0.0546$ \***<br>**SE=0.0267**<br>**p=0.0401** | N = 2,920<br>(14.6%)<br>univariate:<br>**$\beta = 0.0630$ \*\***<br>**SE=0.0205**<br>**p=0.0021**<br>with<br>covariates:<br>$\beta = 0.0403$ †<br>SE=0.0209<br>p=0.0540 | N = 1,381<br>(6.9%)<br>univariate:<br>**$\beta = 0.0908$ \*\***<br>**SE=0.0284**<br>**p=0.0014**<br>with<br>covariates:<br>**$\beta = 0.0594$ \***<br>**SE=0.0285**<br>**p=0.0375** |

| DEPRESSION<br>MEASURE | Never<br>depressed<br>(reference<br>group) | Single<br>Episode<br>Depression | Recurrent<br>Depression |
| --- | --- | --- | --- |
| CIDI-sf plus<br>major role<br>impairment as<br>described by Cai<br>et al. (2020) -<br>Online follow-up<br>Total N = 12,726 | N = 9,472<br>(74.4%)<br>Reference<br>Group | N = 1,256<br>(9.9%)<br>univariate:<br>$\beta = 0.0534$ †<br>SE=0.0298<br>p=0.0726<br>with<br>covariates:<br>$\beta = 0.0436$<br>SE=0.0299<br>p=0.1450 | N = 1,998<br>(15.7%)<br>univariate:<br>$\beta = 0.0302$<br>SE=0.0244<br>p=0.2149<br>with<br>covariates:<br>$\beta = 0.0072$<br>SE=0.0251<br>p=0.7730 |

